# Sickle Cell Disease and Uterine Fibroids: Evaluation of the Prevalence of Fibroids across Sickle Cell Genotypes

**DOI:** 10.1101/2024.03.07.24303932

**Authors:** Jonathan G. Martin, Alexis M. Medema, Blaire K. Rikard, Gabrielle van den Hoek, Miriam Chisholm

## Abstract

**Introduction:** Uterine fibroids are known to affect >80% of premenopausal American women of African descent, and sickle cell disease is known to disproportionately affect people of varying geographical ancestries, particularly those of sub-Saharan African descent. However, previous studies have suggested the two pathologies less frequently co-occur. This study aims to evaluate the prevalence of uterine fibroids in patients with sickle cell disease across a large metropolitan area in the United States.

**Methods:** African American women with sickle cell disease (including HbSS, HbSC, and sickle cell trait genotypes) underwent pelvic imaging (CT/MRI/ultrasound) between February 2011 and August 2018 at two large hospital systems within a single academic institution. Based on retrospective review, the prevalence of uterine fibroids among this cohort was analyzed and compared to published data of fibroid prevalence amongst African American patients without sickle cell disease.

**Results:** Prior data estimates that the prevalence of uterine fibroids in African American women is about 32 to 40% for those aged 32 to 35 years and up to >80% in premenopausal African American women overall. When compared to the expected prevalence in this cohort, with a median age of 31 years, women with HbSS or HbSC sickle cell disease had a significantly decreased prevalence of uterine fibroids (9.6 to 10.3%), while those with sickle cell trait reflected a prevalence (44.4%) like that of the general population.

**Conclusion:** There was a significantly lower prevalence of uterine fibroids in premenopausal American women of African heritage with sickle cell disease in the study cohort when compared to premenopausal American women of African heritage in the general population. This suggests a higher threshold to ascribe dysfunctional uterine bleeding in premenopausal African-American women with sickle cell disease to uterine fibroids, and a lower threshold to pursue an alternative diagnosis.

## INTRODUCTION

Sickle cell disease (SCD) refers to a collection of genetic blood disorders that affect an estimated 100,000 Americans and occur in approximately one of every 365 Black or African American births.^1,2^ The pathophysiology stems from a single amino acid substitution in the gene encoding the beta-globin subunit, causing red blood cells to sickle into a rigid form that is prone to vaso-occlusion, and thus leads to a range of clinical complications.^2^Forms of SCD range from the most common and most severe sickle cell anemia caused by homozygous hemoglobin SS (HbSS), to the heterozygous hemoglobin SC (HbSC), and to SCD trait, based on the relative percentage of mutated and wildtype hemoglobin. In the clinical context, SCD patients often interface with interventional radiology (IR) in the setting of obtaining vascular access or addressing vascular access complications.^3,4^

Uterine leiomyomas, commonly referred to as uterine fibroids (UF), are benign, smooth muscle tumors with an estimated incidence in African American women of up to 40% by age 35 and >80% by age 50.^5,6^Clinically, UF may present with abnormal uterine bleeding, pelvic pain, and reproductive problems, or they may be asymptomatic.^7^Traditionally, treatment options for UF included gynecologic intervention with hysterectomy or myomectomy. However, the American College of Obstetrics and Gynecology (ACOG) now also recommends uterine artery embolization (UAE) for patients with symptomatic fibroids who desire uterine preservation. ^8^

Performed by IR, UAE blocks the arterial supply to UFs and has been demonstrated to be effective in symptomatic management.^9,10^

Prior work has suggested a protective effect of SCD against the development of UF,^11^prompting further investigation. In this study, we investigated the relationship between SCD and UFs in a large metropolitan city with a 54% Black or African American demographic^12^ in order to better understand the frequency at which the two pathologies co-occur.

## MATERIAL AND METHODS

In this retrospective study, patients treated at two large metropolitan medical centers under one academic institution from February 2011 to August 2018 were analyzed. Inclusion criteria were age ≥18 years, prior imaging of the pelvis (including CT, MRI, and ultrasound), and a documented history of sickle cell disease (including HbSS, HbSC, and sickle cell trait genotypes). Patients with a documented history of sickle cell disease but of unknown etiology were recorded as having SCD not otherwise specified (HbNOS).

The institution’s electronic health record was searched using the key terms: “leiomyoma,” “fibroids,” and “sickle,” identifying only the patients whose final radiology reports contained these terms. At this point, the patient cohort was further narrowed into “sickle *and* fibroid,” “sickle *and* fibroids,” sickle *and* leiomyoma,” and “sickle *and* uterus,” and “sickle *and* uterus *not* absent,” groups. This data set was manually analyzed and duplicated patients and studies were removed. The electric medical records of patients with SCD and UF were then analyzed for age at most recent evaluation, race, gender, sex, admission dates (if relevant), discharge and visit diagnoses, clinical exam notes, operative notes, hematology notes, imaging reports, and pathology reports.

Confidence intervals for proportions were based on the binomial distribution. Proportions were compared using chi-squared tests, calculated using JMP^®^ Pro version 13 (SAS Institute Inc., Cary, NC) with a significance level p<0.05. This study was approved by the institutional review board at the Emory University School of Medicine.

## RESULTS

Initial query identified 19,423 imaging studies with the key word “fibroids,” 3253 studies with the key word “sickle,” and 1590 studies with the key word “leiomyoma.” These results were then narrowed with more exact search terms, revealing 14 “sickle *and* fibroid” studies, 34 “sickle *and* fibroids” studies, 4 “sickle *and* leiomyoma” studies, 705 “sickle *and* uterus” studies, and 477 “sickle *and* uterus *not* absent” studies. After manually reviewing these records for accuracy and removing duplicates, the total number of SCD patients with pelvic imaging for hospitals one and two were 160 and 132, respectively (Figure 1). The final patient demographic data is presented in Table 1.

**Table 1.** Patient demographics and characteristics.

|  | **Hospital #1**<br>N = 160 | **Hospital #2**<br>N = 132 | **Total**<br>N=192 |
| --- | --- | --- | --- |
| **Demographics** |  |  |  |
| Median age (range) | 33 years<br>(19–78) | 29 years<br>(18–69) | 31 years<br>(18–78) |
| African-American/Black | 157 (98.7%) | 121 (91.7%) | 278 (95.2%) |
| **Sickle cell disease genotypes** |  |  |  |
| HbSS | 114 (71.3%) | 91 (68.9%) | 205 (70.2%) |
| HbSC | 8 (5.6%) | 27 (20.5%) | 35 (12%) |
| HbNOS | 30 (18.9%) | 4 (3.0%) | 34 (11.6%) |
| SCD Trait | 8 (5.6%) | 10 (7.6%) | 18 (6.2%) |
| **Patients with UFs (% of genotype)** |  |  |  |
| HbSS | 9 (7.9%) | 11 (12.1%) | 20 (9.8%) |
| HbSC | 1 (12.5%) | 2 (7.4%) | 3 (8.6%) |
| HbNOS | 2 (6.7%) | 0 (0.0%) | 2 (5.9%) |
| SCD Trait | 4 (50%) | 4 (40%) | 8 (44.4%) |

**Figure 1.**
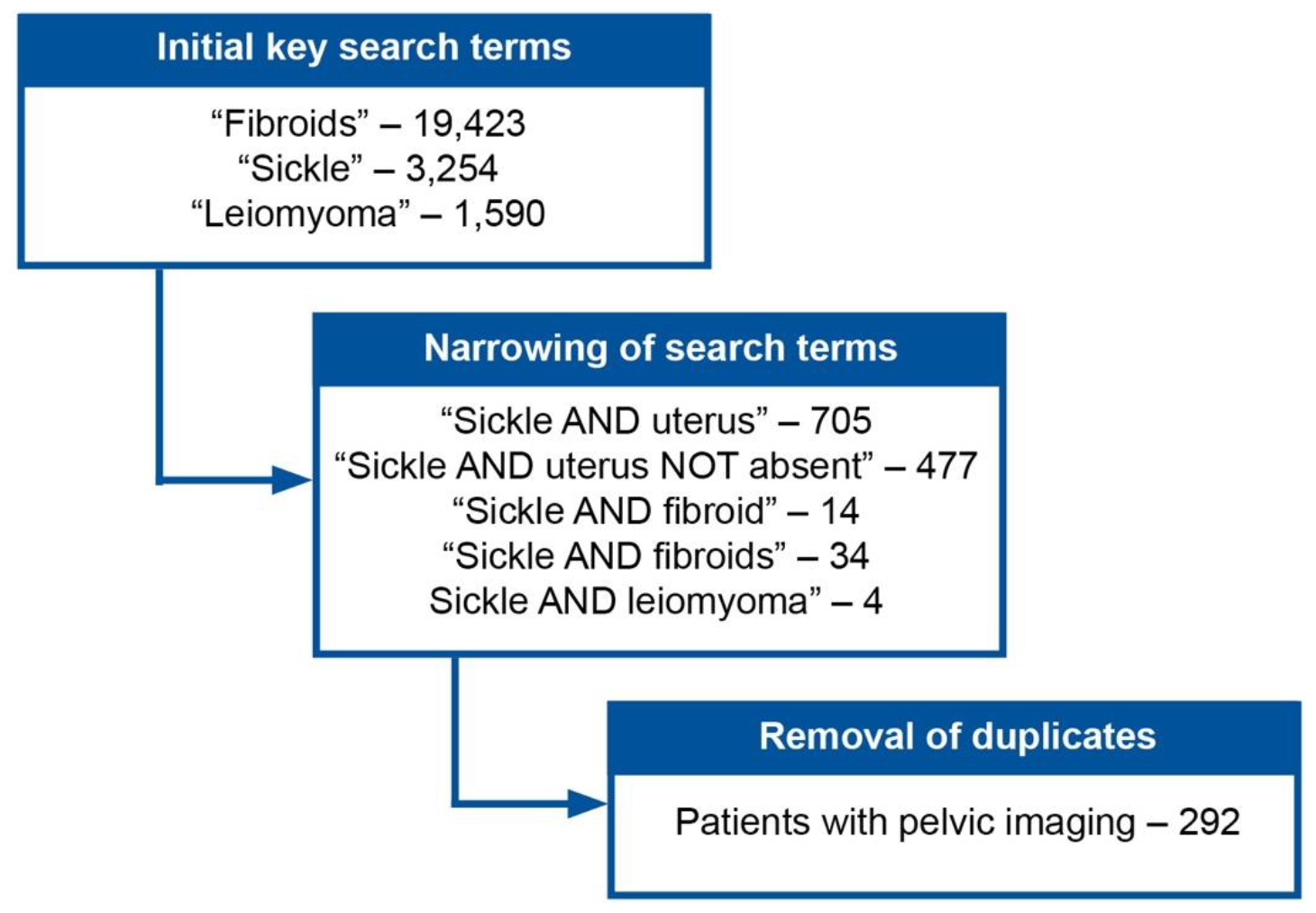
Electronic health record search criteria and refined results (patients identified per search term).

Out of an overall 292 SCD patients, 33 (11.3%, 95% CI=7.9–15.5%) had fibroids as identified on imaging. Of these 33 patients, the median age was 31 years. Among 18 patients with sickle cell trait, 8 (44.4%, 95% CI=21.5–69.2%) had fibroids. Of 240 patients with documented SCD due to either HbSS or HbSC, 23 women had UFs (9.6%, 95% CI=6.1–14.0%) which was significantly different when compared to those with sickle cell trait, (p<0.001).

For 34 HbNOS patients whose SCD type was not definitely identified, 2 (5.9%) had UFs. Even if it is assumed that only the 2 patients with UF were HbSS/HbSC, the adjusted prevalence (10.3%, 95% CI=6.8–14.9%) remains significantly below that expected for an age-adjusted population.

The prevalence of uterine fibroids by sickle cell genotype remained similar across location when patients were further stratified by their primary treating hospital (Figure 2).

**Figure 2.**
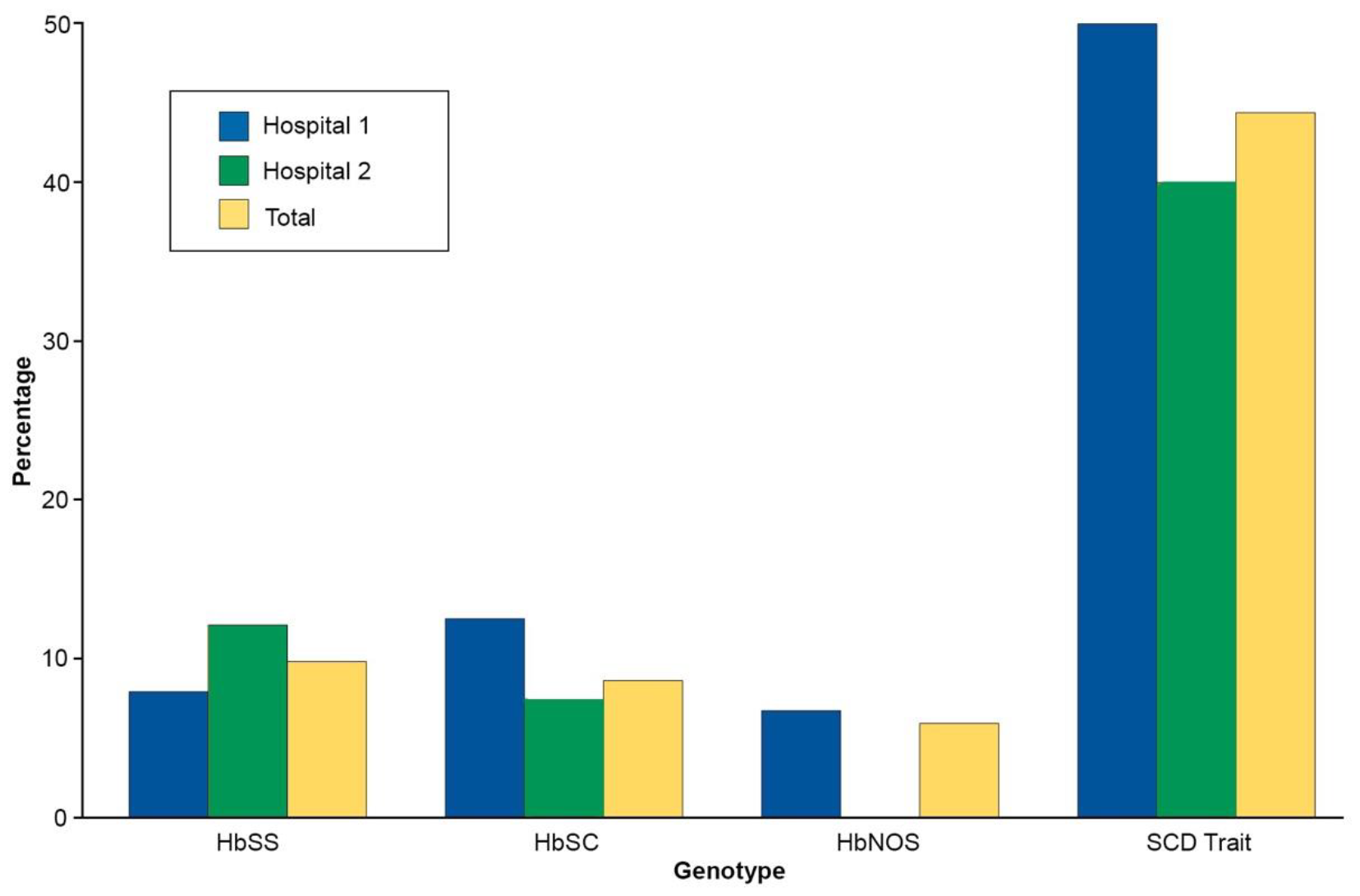
Percentage of SCD patients, by genotype, with uterine fibroids confirmed by imaging.

## DISCUSSION

In premenopausal African American women without SCD, the prevalence of UF ranged from 10% for those aged 23–25 years, to 32% for those aged 32–35 years, and up to 87% overall.^5,6^ Given the median age of 31 years in the present study, our prevalence of fibroids of 9.6–10.3% for those with sickle cell disease was significantly lower than that expected for the general population of premenopausal African American women. These findings are consistent with prior work demonstrating a prevalence of uterine fibroids of 0% in patients aged 20–29 years and of 13.8% in patients aged 30–39 years in patients with sickle cell disease.^11^ The clinical impact of such an association, if confirmed by additional investigation, would be to suggest that a higher threshold is needed to ascribe dysfunctional uterine bleeding in premenopausal African-American women with sickle cell disease to uterine fibroids, and that in these individuals a lower threshold should be held to pursue an alternative diagnosis.

Interestingly, the decreased prevalence of UFs in patients with HbSS and HbSC disease stands in contrast with the 44.4% prevalence seen in patients with sickle cell trait, which more closely mirrors that of the general population.^5,6^ This contrast suggests that sickle cell trait may not confer a similar protective effect against the development of fibroids. Traditionally, sickle cell trait has been considered a benign condition, with patients who inherit only one copy of a mutated beta-globin gene typically spared from the vaso-occlusive crises characteristic of sickle cell anemia.^2,13^ As such, the impaired oxygen supply seen in HbSS and HbSC patients but not sickle cell trait patients, when combined with the baseline hypoxic environment of UF development, may play a role in fibroid cell survival, as suggested by prior work.^11^

This study has many limitations, including its retrospective design, limited geographic area, and referral selection bias. In a 2013 study by Stewart et al., African American women were significantly more likely to seek treatment for UF symptoms later than white women,^14^ and thus reporting bias is a limitation. Although this study controlled for duplicated patients who visited different hospitals within the same metropolis, women may have sought UF treatment at other institutions, which may have reduced the population observed. Additionally, by identifying the patient cohort via key terms within imaging reports, it is possible that patients with SCD may have been inadvertently left out of the study due to absence of those selected terms in their reports. Similarly, in the current study, CT, MRI, and ultrasound imaging were considered adequate for the diagnosis of leiomyoma. However, prior research has demonstrated that MRI may be superior for the diagnosis of numerous osseous, soft tissue, and solid organ pathologies,^15-17^ including uterine fibroids and adenomyosis.^18,19^ There may be other confounding variables inherent in the two different patient populations that were not readily identifiable by retrospective review.

## CONCLUSIONS

The relationship between SCD and UFs is complex, but the significantly decreased prevalence of UFs in African American women with SCD compared to previously published data suggests an inherent physiologic mechanism in SCD that reduces the risk of UFs as tumors that depend on a robust blood supply. These preliminary findings recommend additional research involving a larger and more geographically diverse population that may confirm this relationship, as well as work to clarify the biochemical etiologies underlying this association.

## Data Availability

All data produced in the present study are available upon reasonable request to the authors.

## Notes

### Competing Interest Statement

The authors have declared no competing interest.

### Funding Statement

This study did not receive any funding.

### Author Declarations

IRB of Emory University School of Medicine gave ethical approval for this work.

